# A multiplex Mtb-specific FluoroSpot assay measuring IFNγ, TNF, and IL2-secreting cells can improve accuracy and differentiation across the tuberculosis spectrum

**DOI:** 10.1101/2025.05.05.25326945

**Authors:** Elin Folkesson, Fariba Foroogh, Vera Kjellgren, Mathilda Jakobsson, Lisann Grunewald, Jens Hellberg, Carolina Sousa Silva, Maia S. Gower, Linn Kleberg, Hans Grönlund, Margarida Correia-Neves, Bartek Makower, Gunilla Källenius, Judith Bruchfeld, Christopher Sundling

## Abstract

Tuberculosis (TB) remains a major global health challenge, with approximately 25% of the global population estimated to have been infected with the causative pathogen *Mycobacterium tuberculosis (Mtb)*. Current diagnostic methods based on Interferon-gamma (IFNγ) release assays (IGRA) have limitations in detecting infection and cannot distinguish those with TB infection from those with disease. In this study, we evaluated a multiplex FluoroSpot assay measuring IFNγ, TNF, and IL2-secreting cells in response to the *Mtb* antigens ESAT-6, CFP-10, and EspC. The assay was tested on peripheral blood mononuclear cells from individuals with TB disease (n=24), TB infection (n=63), and IGRA-negative controls (n=27). Results indicate that triple cytokine-secreting cells (IFNγ/IL2/TNF) could detect *Mtb*-specific immune responses with higher sensitivity and specificity compared to commercially available IGRA methods. Furthermore, we identified distinct cytokine profiles associated with different stages of TB infection and disease. The study suggests that this multiplex assay could improve current TB diagnostics while also improving our understanding of *Mtb*-specific T cell responses associated with recent or remote TB infection and disease. Further studies are now needed to validate these findings in larger cohorts, including individuals with immunosuppressive conditions.

## Introduction

Tuberculosis (TB) is a significant public health issue with an estimated 10.8 million cases of TB disease (TBD) in 2023, resulting in 1.25 million deaths, thus making it the leading cause of mortality caused by a single infectious agent [1]. The persistently high incidence is the result of ongoing transmission, but also due to the capacity of *Mycobacterium tuberculosis* (*Mtb*) to establish a TB infection (TBI) that will most often pass unnoticed but after a variable length of time can resurge and cause TBD. Current estimations infer that approximately 25% of the world’s population have TBI, defined as reactivity for Mtb in immunological tests without clinical or radiological evidence of TB disease [2]. This reservoir of possible future incident TB cases needs to be addressed to achieve the end TB 2035 goals [3], aiming to reduce TB incidence by 90% and TB mortality by 95% compared to 2015 numbers [4].

A historic simplified notion of *Mtb* as either causing TBD (symptomatic disease) or TBI (persistent non-symptomatic infection) has been replaced with the understanding that these are different states of a disease spectrum [5–7]. Following exposure to *Mtb* through inhaled aerosolized droplets the immune response can either clear the bacteria or, failing to do so, an infection will be established. When the adaptive immune system becomes involved, the differentiation of *Mtb* specific T-cells will usually be associated with bacterial containment. The balance between *Mtb* specific immunological control and mycobacterial survival will determine the outcome in the individual patient, with around 5 % progressing to TBD within the first couple of years after infection and an estimated 10% lifetime risk for TBD overall [5,7]. It is not known which proportion of individuals will eventually clear the infection over time though some argue that lifelong infection is less common than previously believed [8], and currently there are no methods to detect true persistence of viable *Mtb* [9]. Instead, the diagnosis of TBI is based on indirect immunological methods, detecting a specific T-cell mediated immune response to *Mtb* antigens, through Tuberculin Skin Tests (TST) or Interferon-gamma release assays (IGRA) in blood [10]. Although the more recent IGRA’s, Quantiferon-Gold in Tube (QFT-GIT, from Qiagen) and T-SPOT.TB (from Oxford Immunotech), have an advantage over TST in being more specific [11], neither can differentiate between TBD and TBI, nor can they predict disease progression or clearance of *Mtb* [12][13].

In non-immunosuppressed adults the estimated sensitivity for TBI is around 80% for TST and QFT-GIT and 90% for T-SPOT.*TB* although a reference method is not available for direct comparison [14,15]. Sensitivity is lower in immunosuppressed patients due to HIV infection or other immunosuppressive conditions and treatments [16–18], in children [19,20] and with higher rates of indeterminant results [21,22]. These limitations cause significant clinical dilemmas as these groups are also at higher risk for progression to TBD [22]. Furthermore, the positive predictive value for developing TBD is low for all tests, with T-SPOT.TB reaching at most 4.2% [23–25]. Testing for TBI and providing preventive TB treatment (PTT) is recommended by WHO to persons living with HIV [26], household contacts of individuals with positive pulmonary TB, and in anticipation of certain immunosuppressive treatments [27]. In order to scale up PTT, more accurate testing for TBI is warranted, including tests that can differentiate recent and remote TBI and identify individuals with high risk of TB disease progression [12,28,29], as well as tests with high sensitivity in immunosuppressed individuals [18].

The WHO endorsed [10] IGRA tests use *Mtb* antigens ESAT-6 and CFP-10 to stimulate T-cells in whole blood (QTF-Plus and WANTAI TB IGRA) or isolated peripheral blood mononuclear cells (PBMCs) (T-SPOT.*TB*) and measures interferon-γ (IFNγ) after 24 hours incubation by ELISA or EliSpot respectively. In recent years other methods, that are more suitable in low resource settings and for screening purposes, have been developed, while still using the same *Mtb* antigen/IFNγ combination [24,30]. Possible ways to improve these assays could be adding other *Mtb* antigens to elicit stronger or differential responses according to *Mtb* infection stage, or measuring combinations of cytokines [31,32]. The so called *Mtb* latency antigens, all located within the Dormancy survival regulon (DosR) have shown some potential to differentiate TBI from TBD [32] and EspC, a protein involved in secretion of *Mtb* virulence factors, is demonstrated to elicit strong immune responses, improving sensitivity for TBD when added to ESAT-6 and CFP-10 [33].

There is also increasing support for using non-IFNγ *Mtb*-specific immune responses, with the cytokines tumor necrosis factor (TNF), interleukin (IL) 2 and IFNγ-inducible protein 10 (IP-10) among the most widely investigated. For example, secretion of TNF and IL2 in response to *Mtb*-specific (ESAT-6 and CFP-10) stimulation in extensively *Mtb*-exposed IGRA negative individuals have been described, possibly indicating a protective TB immune response [34,35]. Furthermore, so called in-vitro expressed TB antigens (IVE-TB antigens) identified through transcriptomic analysis, have been shown to elicit non-IFNγ responses, including TNF, IP-10 and IL-17 [36,37]. IP-10, which is released following IFNγ stimulation, has been suggested as a more sensitive marker due to amplification of the IFNγ signal [38–40]. A recent study found sensitivity of an assay using IP-10 and IL-2 following ESAT-6/CFP-10 stimulation equal to IFNγ in detecting TBI [41]. According to reviews of TBI diagnostics [24,30], IP-10 is included in a new immunoassay for TBI currently in development (rBiopharm, Darmstadt, Germany).

Identifying polyfunctional T-cells with distinct cytokine-patterns may improve diagnostic sensitivity, and in addition potentially inform about different *Mtb* infection stages. A promising approach is the FluoroSpot technology, which builds on the ELISpot method used in T-SPOT.*TB* but uses fluorescence labelled cytokine-specific antibodies to simultaneously detect multiple cytokines [42]. In our laboratory we have previously developed a multiplex FluoroSpot that could simultaneously assess four separate B cell effector responses at a single-cell level [43–45]. Building on this experience we are now, in collaboration with the Swedish biotech company Mabtech AB, optimizing an *Mtb*-specific FluoroSpot™ (Mabtech) assay that can also detect up to four cytokines per cell. In addition to enumerating antigen-specific cells it offers a semi-quantitative measure of the secreted cytokines by evaluating the volume of the spots [46].

In this FluoroSpot™ assay we used the *Mtb* antigens ESAT-6, CFP-10 and EspC for PBMC stimulation and the cytokine panel IFNγ/IL2/TNF to evaluate memory T cell responses. We hypothesize that the multiplex cytokine detection would allow for increased sensitivity with potential to differentiate disease states and T-cell function better than IFNγ alone. Moreover, the addition of EspC could offer an increased sensitivity as additional T cells could be detected. The main aim of this study was to evaluate this FluoroSpot method with regards to its sensitivity and specificity in TBD and TBI, and whether different stages of *Mtb* infection may be identified by patterns observed in terms of frequency, magnitude and combination of single/double/triple cytokine producing cells.

## Results

### Study participants

A total of 115 study participants were recruited for the study, falling into three distinct groups: individuals with microbiologically verified TBD (n=24), TBI (n=63), and non-TB IGRA negative controls (n=27). The TBD group consisted of individuals with pulmonary TB (n=13) and extrapulmonary TB (n=11), with culture-verification in 23 (96%) individuals in total. The study groups had similar gender distribution, age, co-morbidities and immunosuppression (*Table 1*). Most individuals in both TB groups were non-Swedish born, a majority from TB high-endemic countries (>100/100 000, [47]), while fewer (37%) IGRA negative controls were from TB high-endemic countries. Individuals with TBD had significantly higher C-reactive protein (CRP) and erythrocyte sedimentation rate (ESR), and lower hemoglobin and albumin levels compared to those with TBI (*Table 1*).

**Table 1.**
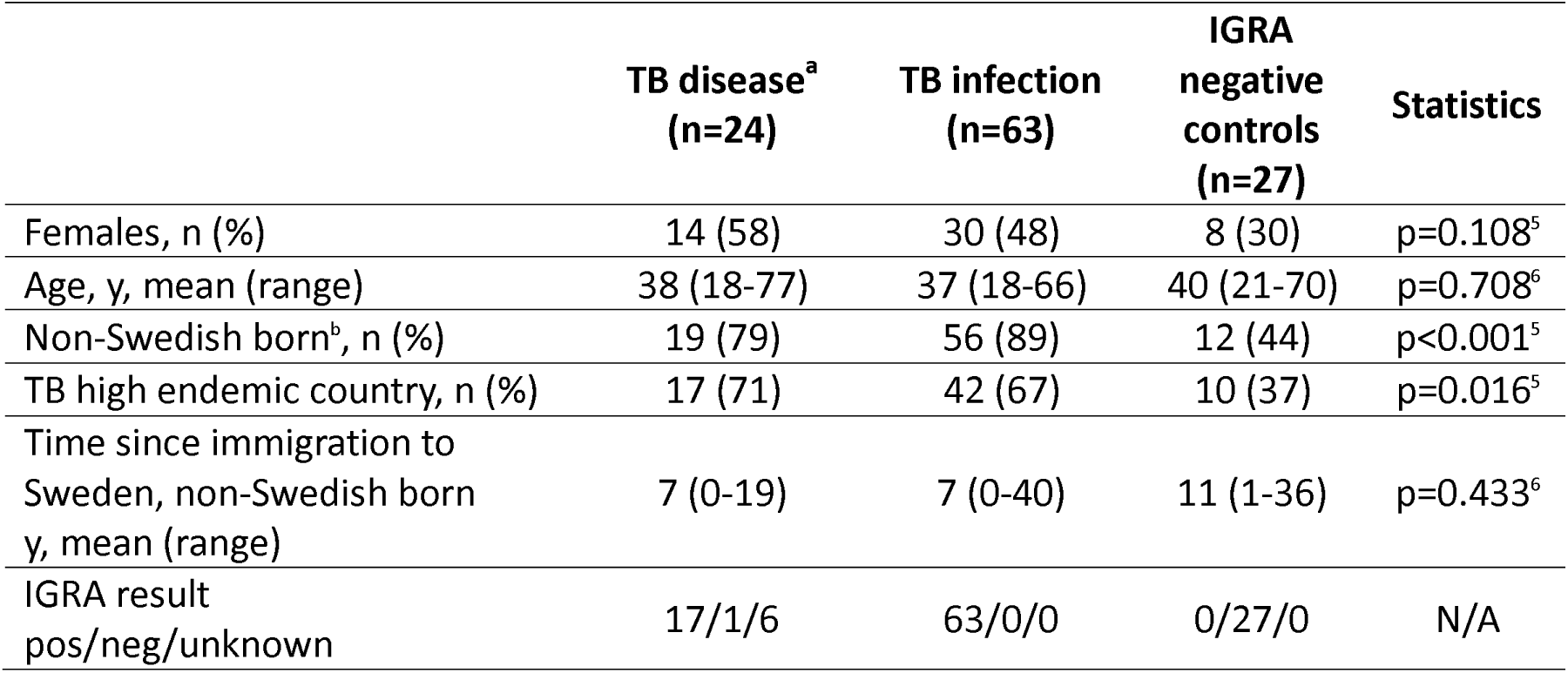
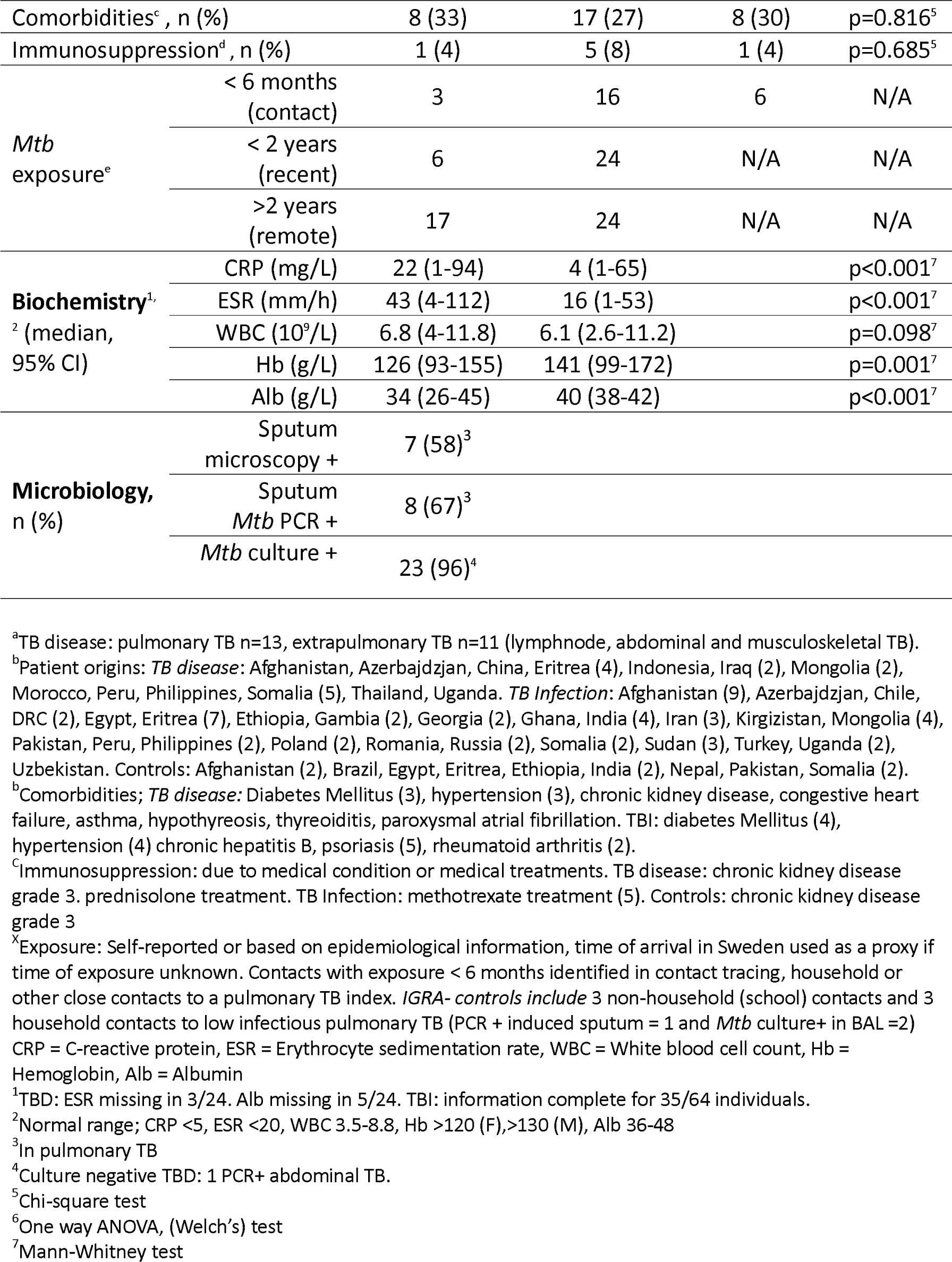
Clinical characteristics of the patient cohort tested with the FluoroSpot assay.

|  | TB disease<sup>a</sup><br>(n=24) | TB infection<br>(n=63) | IGRA<br>negative<br>controls<br>(n=27) | Statistics |
| --- | --- | --- | --- | --- |
| Females, n (%) | 14 (58) | 30 (48) | 8 (30) | p=0.108<sup>5</sup> |
| Age, y, mean (range) | 38 (18-77) | 37 (18-66) | 40 (21-70) | p=0.708<sup>6</sup> |
| Non-Swedish born<sup>b</sup>, n (%) | 19 (79) | 56 (89) | 12 (44) | p<0.001<sup>5</sup> |
| TB high endemic country, n (%) | 17 (71) | 42 (67) | 10 (37) | p=0.016<sup>5</sup> |
| Time since immigration to<br>Sweden, non-Swedish born<br>y, mean (range) | 7 (0-19) | 7 (0-40) | 11 (1-36) | p=0.433<sup>6</sup> |
| IGRA result<br>pos/neg/unknown | 17/1/6 | 63/0/0 | 0/27/0 | N/A |

|  |  |  |  |  |  |
| --- | --- | --- | --- | --- | --- |
| Comorbidities<sup>c</sup>, n (%) |  | 8 (33) | 17 (27) | 8 (30) | p=0.816<sup>5</sup> |
| Immunosuppression<sup>d</sup>, n (%) |  | 1 (4) | 5 (8) | 1 (4) | p=0.685<sup>5</sup> |
| *Mtb* exposure<sup>e</sup> | < 6 months (contact) | 3 | 16 | 6 | N/A |
|  | < 2 years (recent) | 6 | 24 | N/A | N/A |
|  | >2 years (remote) | 17 | 24 | N/A | N/A |
| **Biochemistry**<sup>1</sup><br><sup>2</sup> (median, 95% CI) | CRP (mg/L) | 22 (1-94) | 4 (1-65) |  | p<0.001<sup>7</sup> |
|  | ESR (mm/h) | 43 (4-112) | 16 (1-53) |  | p<0.001<sup>7</sup> |
|  | WBC (10<sup>9</sup>/L) | 6.8 (4-11.8) | 6.1 (2.6-11.2) |  | p=0.098<sup>7</sup> |
|  | Hb (g/L) | 126 (93-155) | 141 (99-172) |  | p=0.001<sup>7</sup> |
|  | Alb (g/L) | 34 (26-45) | 40 (38-42) |  | p<0.001<sup>7</sup> |
| **Microbiology,**<br>n (%) | Sputum microscopy + | 7 (58)<sup>3</sup> |  |  |  |
|  | Sputum *Mtb* PCR + | 8 (67)<sup>3</sup> |  |  |  |
|  | *Mtb* culture + | 23 (96)<sup>4</sup> |  |  |  |

<sup>a</sup>TB disease: pulmonary TB n=13, extrapulmonary TB n=11 (lymphnode, abdominal and musculoskeletal TB).
<sup>b</sup>Patient origins: *TB disease*: Afghanistan, Azerbajdzjan, China, Eritrea (4), Indonesia, Iraq (2), Mongolia (2), Morocco, Peru, Philippines, Somalia (5), Thailand, Uganda. *TB Infection*: Afghanistan (9), Azerbajdzjan, Chile, DRC (2), Egypt, Eritrea (7), Ethiopia, Gambia (2), Georgia (2), Ghana, India (4), Iran (3), Kirgizistan, Mongolia (4), Pakistan, Peru, Philippines (2), Poland (2), Romania, Russia (2), Somalia (2), Sudan (3), Turkey, Uganda (2), Uzbekistan. Controls: Afghanistan (2), Brazil, Egypt, Eritrea, Ethiopia, India (2), Nepal, Pakistan, Somalia (2).
<sup>c</sup>Comorbidities; *TB disease*: Diabetes Mellitus (3), hypertension (3), chronic kidney disease, congestive heart failure, asthma, hypothyreosis, thyroiditis, paroxysmal atrial fibrillation. TBi: diabetes Mellitus (4), hypertension (4) chronic hepatitis B, psoriasis (5), rheumatoid arthritis (2).
<sup>d</sup>Immunosuppression: due to medical condition or medical treatments. TB disease: chronic kidney disease grade 3. prednisolone treatment. TB Infection: methotrexate treatment (5). Controls: chronic kidney disease grade 3
<sup>e</sup>Exposure: Self-reported or based on epidemiological information, time of arrival in Sweden used as a proxy if time of exposure unknown. Contacts with exposure < 6 months identified in contact tracing, household or other close contacts to a pulmonary TB index. *IGRA- controls include* 3 non-household (school) contacts and 3 household contacts to low infectious pulmonary TB (PCR + induced sputum = 1 and *Mtb* culture+ in BAL =2)
CRP = C-reactive protein, ESR = Erythrocyte sedimentation rate, WBC = White blood cell count, Hb = Hemoglobin, Alb = Albumin
<sup>1</sup>TBD: ESR missing in 3/24. Alb missing in 5/24. TBi: information complete for 35/64 individuals.
<sup>2</sup>Normal range; CRP <5, ESR <20, WBC 3.5-8.8, Hb >120 (F), >130 (M), Alb 36-48
<sup>3</sup>In pulmonary TB
<sup>4</sup>Culture negative TBD: 1 PCR+ abdominal TB.
<sup>5</sup>Chi-square test
<sup>6</sup>One way ANOVA, (Welch's) test
<sup>7</sup>Mann-Whitney test

### Comparable detection of Mtb-specific IFNγ-secreting cells with ELISpot and multiplex FluoroSpot

The transition from an ELISpot system (used in the T-Spot.TB IGRA assay) to a multiplex fluorescent detection of cytokine secretion could potentially impact the detection sensitivity for cytokine secretion. Therefore, we first measured the number of IFNγ secreting cells, presented as spot forming units (SFU), from a subset of donors with TBD (n=10), TBI (n=14), or IGRA negative controls (n=7) using both IFNγ/IL2/TNF multiplex FluoroSpot and IFNγ ELISpot. PBMCs were stimulated overnight with the *Mtb*-specific proteins ESAT-6, CFP-10, and EspC, targeted toward MHC class II presentation and memory CD4 T cells (**Figure 1A**). Comparing the IFNγ detection only, both methods detected IFNγ producing cells to a comparable extent (**Figure 1B**) with the number of SFUs being highly correlated (r^2^=0.72, p<0.001) (**Figure 1C**).

**Figure 1.**
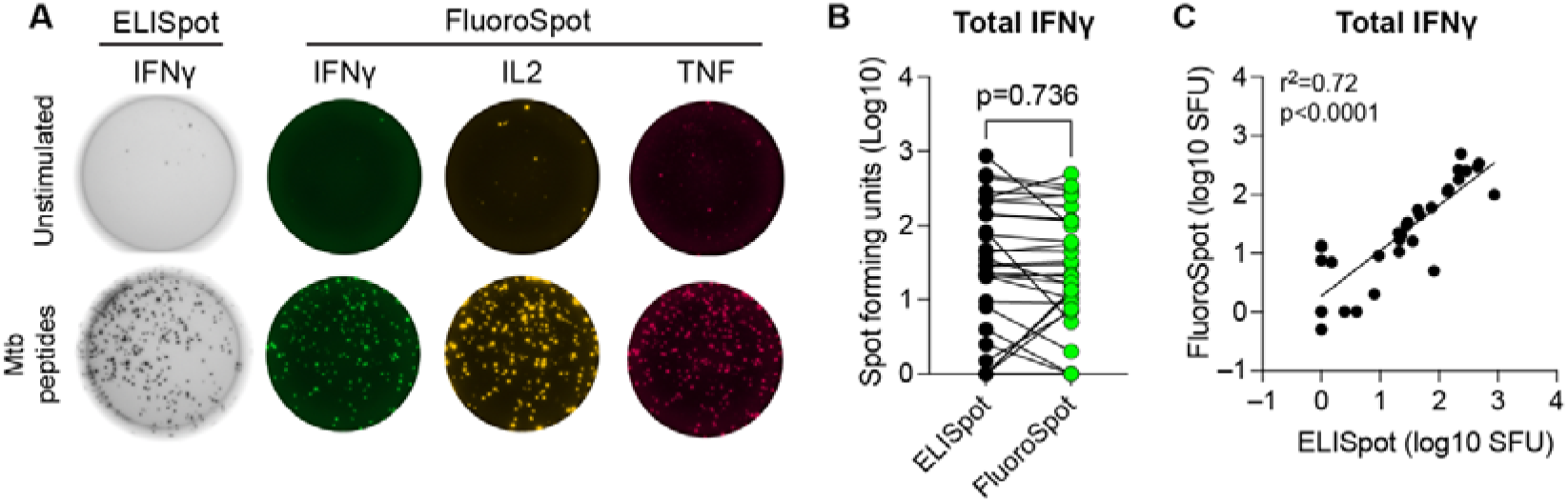
IFNγ ELISpot and FluoroSpot results show high agreement in spot forming units upon stimulation with ESAT-6, CFP-10, and EspC. (**A**) Representative wells from FluoroSpot and ELISpot without stimulation (top row) and with overnight stimulation (bottom row) with peptide pools from Mtb proteins ESAT-6, CFP-10, and EspC. In the bottom row each colored spot equals one SFU for the respective cytokine measured (IL2, TNF and IFN*γ*). (**B**) IFN*γ*-specific SFU from paired samples analyzed by both ELISpot and multiplex FluoroSpot. Statistics were evaluated by a paired t-test. (**C**) Pearson correlation (r^2^) of log10-transformed IFNγ SFU for FluoroSpot and ELISpot after stimulation. Each dot represents one donor (n=31). **Abbreviations:** spot forming unit (SFU)

### Lower background when using polyfunctional cytokine secretion to detect Mtb infection

We next assessed the secretion of the different cytokine combinations with and without stimulation with *Mtb* antigens. *Mtb*-peptide stimulation led to an increased number of SFU for all IFNγ and IL2 cytokine combinations in both TBD and TBI groups. (**Figure 2A**). Single positive IL2 SFUs were also detectable for IGRA negative controls, indicating some non-specific IL2-secretion. TNF was the cytokine secreted by the largest number of cells (**Figure 2A**), however, the secretion was equally high in both *Mtb* stimulated and unstimulated wells and largely coming from TNF single-secreting cells. This suggests that TNF single-secreting cells are not *Mtb*-specific, but rather a result of bystander cell activation. To account for this non-specific cytokine secretion, all subsequent calculations of SFUs were background-subtracted with the SFU from the unstimulated wells.

**Figure 2.**
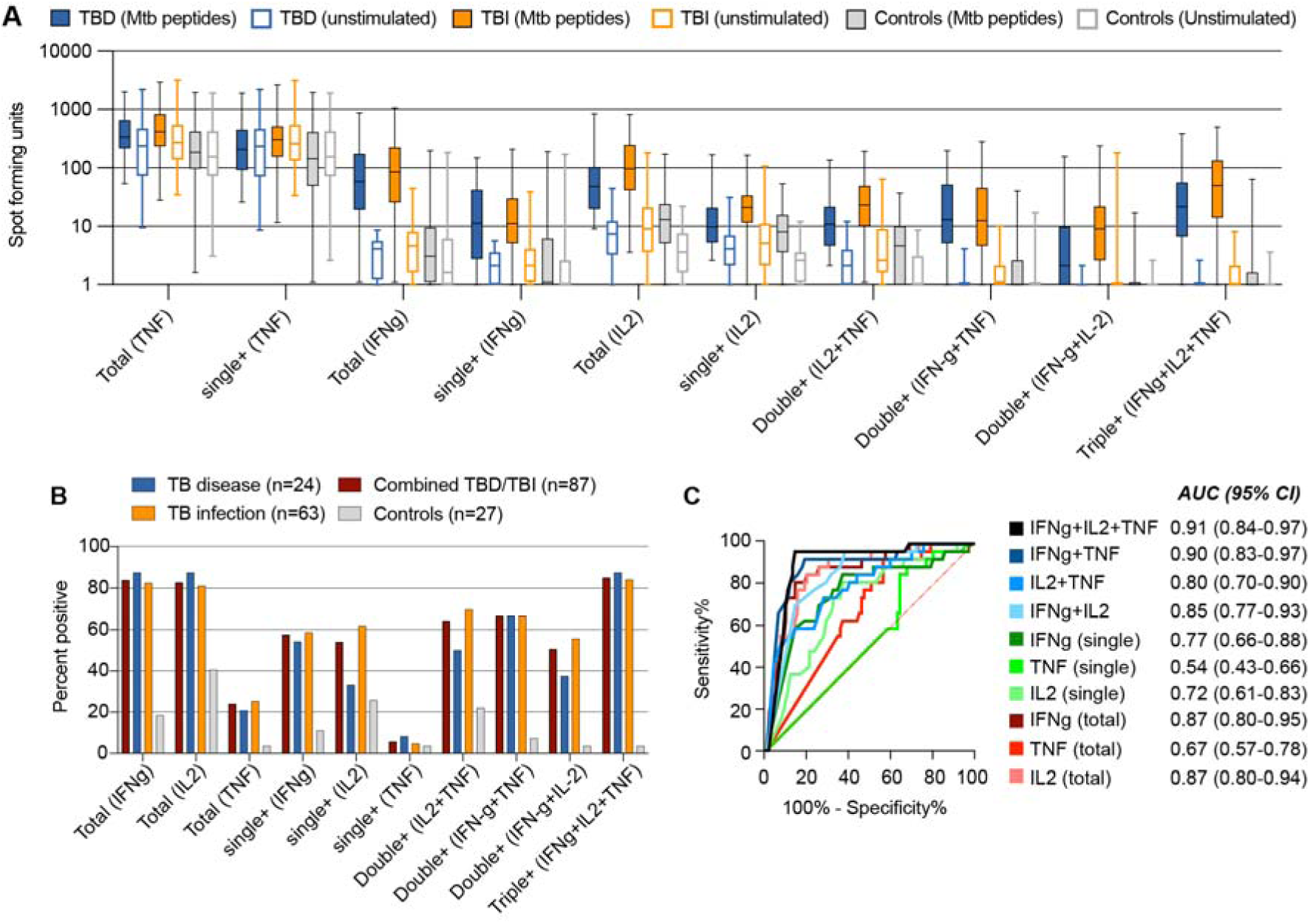
Lower false-positive rates when using IFNγ/IL2/TNF triple positive cells for detection of TB infection and disease. (**A**) The number of SFU in ESAT-6, CFP-10, and EspC stimulated and unstimulated conditions for each group (TBD: TB disease n=24, TBI: TB infection n=63, IGRA negative controls n=27) and IFN*γ*, IL2, TNF cytokine combination. For donors with no detectable cytokines, the SFU was set to 1 to enable visualization on a log scale. (**B**) Frequency of positive donors for individuals with TBD (n=24), TBI (n=63), merged TBD and TBI (n=87), and IGRA negative controls (n=27). The cut-off for positivity was set as [SFU(stimulated) – 2 x SFU(unstimulated)] *≥* 5. Total indicates all cells producing the given cytokine, independent of being single, double, or triple producers. (**C**) ROC analysis with calculation of the AUC +/– 95% confidence interval (CI). TBD and TBI groups combined (n=87) compared to IGRA negative controls (n=27). **Abbreviations:** TBD: TB disease; TBI: TB infection; SFU: spot forming unit; ROC: receiver operating characteristic; AUC: area under the curve; IGRA: interferon gamma release assay.

We calculated the percentage of donors that were positive in the multiplex FluoroSpot assay after stimulation with the *Mtb*-peptides (**Figure 2B**). To ensure that non-specific production of cytokines did not influence the results, we defined the criteria for positivity as: *[SFU(stimulated) – 2 x SFU(unstimulated)]* ≥ 5. We chose five SFUs as the cut-off because the T-Spot.TB IGRA uses <5 SFU as a cut-off to identify negative wells. Triple positive IFNγ/IL2/TNF cells (i.e. all three cytokines detected in the same SFU), total IFNγ, and total IL2 (i.e. any SFU containing IFNγ or IL2 respectively, alone or in combination with any other cytokine) yielded the highest frequency in both the TBD group (88%), in the TBI group (81-84%), and the two TB groups combined (83-85%) (**Figure 2B**). In the IGRA negative control group, triple IFNγ/IL2/TNF secreting cells were detected in only one individual (4%), compared to total IFNγ (19%) and total IL2 (41%), suggesting a higher specificity for the triple IFNγ/IL2/TNF secreting cells. The single IGRA negative donor secreting all three cytokines displayed a strong *Mtb*-specific response (high in *Mtb*-stimulation and low in unstimulated), indicating a false negative result in the clinical QuantiFERON Gold Plus IGRA-test.

For double cytokine-secreting cells, overall lower positivity rates were seen in both TBD and TBI compared to triple IFNγ/IL2/TNF+ and total IFNγ and IL2 (**Figure 2B**). Relatively more individuals with TBI were positive for IL2-combinations (IFNγ/IL2 and IL2/TNF), which was also observed for single IL2+ cells. Single TNF+ stood out by demonstrating low positivity rates in either group, which was primarily due to high background levels indicating non-specific TNF-release.

To measure the overall performance of the different cytokine combinations for detecting an *Mtb*-specific immune response, we combined the TBD and TBI groups and compared to the IGRA negative controls through an area under curve receiver operating characteristic (AUROC) analysis (**Figure 2C**). Based on the number of background-subtracted SFU, AUC was highest for triple IFNγ/IL2/TNF+ cells at 0.91 (95% CI 0.84-0.97), mainly explained by its higher specificity, followed by IFNγ/TNF (AUC=0.9, 95% CI: 0.83-0.97), total IFNγ (AUC=0.87, 95% CI: 0.80-0.95) and total IL2 (AUC=0.87, 95% CI: 0.80-0.94) (**Figure 2C**).

### Increased classification of Mtb infection using IFNγ/IL2/TNF triple-secreting cells compared with total IFNγ

The discovery that the positivity rate for triple IFNγ/IL-2/TNF secreting cells was similar to total IFNγ positive cells in both TB groups (TBD and TBI) but lower in IGRA negative controls, (**Figure 2A**) led us to explore the performance of the triple+ cells compared to standard IFNγ. We next compared the level of SFUs for triple IFNγ/IL2/TNF+ and total IFNγ+ in TBD and TBI versus controls. Both TB groups had higher triple IFNγ/IL2/TNF+ and total IFNγ+ SFUs compared with controls, while no difference was seen between TBD and TBI. As expected, triple IFNγ/IL2/TNF+ SFU were lower compared with total IFNγ+ SFU in all study groups (**Figure 3A**). We next performed separate ROC analysis based on SFU levels in TBD and TBI versus controls, resulting in a higher AUC for the triple IFNγ/IL2/TNF+ in both TBD (AUC 0.91 vs 0.87) and TBI (AUC 0.90 vs 0.87) (**Figure 3B**).

**Figure 3.**
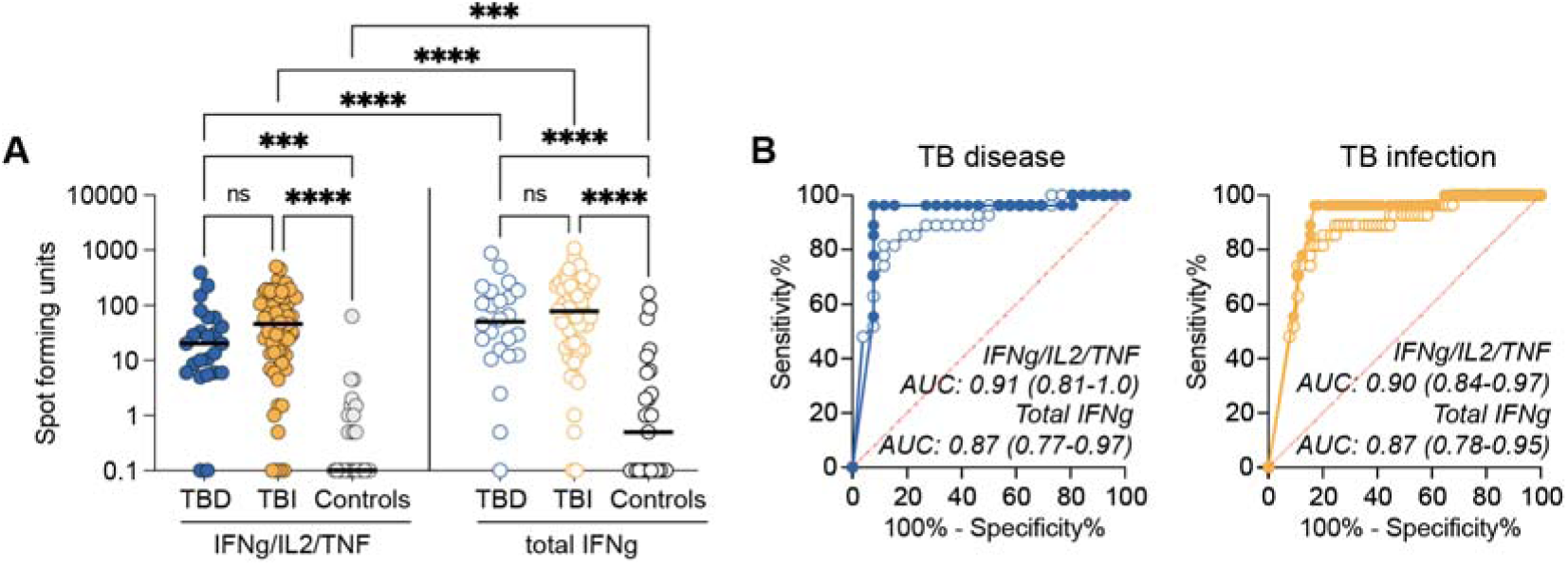
Increased diagnostic performance with the polyfunctional Mtb-specific FluoroSpot assay. (**A**) Background-subtracted SFU for TBD (n=24), TBI (n=63), and IGRA negative controls (n=27). The left panel indicates the SFU count for cells simultaneously secreting IFN*γ*, IL2, and TNF and the right panel indicates the SFU for all cells producing IFN*γ,* whether as single cytokine or in combination with other cytokines. Donors with no detectable cytokine-secreting cells were set to 0.1. Statistics between independent groups were calculated with a Kruskal-Wallis test followed by uncorrected Dunn’s posttest, while comparison between IFN*γ*/IL2/TNF and total IFN*γ* were calculated based on the Wilcoxon matched-pairs test, ns=p>0.05, ***p<0.001, ****p<0.0001. (**B**) AUROC curves for TBD (left panel) and TBI (right panel) comparing IFN*γ*/IL2/TNF+ (Triple; filled circles) and Total IFN*γ*+ (Total; open circles). AUC and 95% confidence intervals are indicated. **Abbreviations:** SFU: spot forming units; TBD: TB disease; TBI: TB infection; IGRA: interferon gamma release assay. AUROC: area under the receiver operating characteristic. AUC: area under the curve

### Diverse functional profiles of cytokine-secreting cells upon peptide stimulation

To investigate if the pattern of cytokine secretion differed between TBD and TBI groups, we calculated the frequency of each cytokine combination out of the *Mtb* specific cytokine-producing cells (**Figure 4**). We excluded single and total TNF from the calculation, considering that our results showed it was mainly derived from bystander activation. In both TB groups (TBD and TBI), cells producing all three cytokines were the most common (median TBD: 37% and TBI: 41%), followed by cells co-producing IFNγ/TNF (median TBD: 20% and TBI: 13%) and IL2/TNF (median TBD: 10% and TBI: 16%) (**Figure 4A**). Directly comparing the frequency of cells secreting the different cytokine combinations in individuals with TBD to those with TBI we found: 1) no difference for triple IFNγ/IL2/TNF+ cells; 2) more double IFNγ/IL2+ and IL2/TNF+ cells in TBI, and; 3) more single IFNγ+ and double IFNγ/TNF+ cells in TBD (**Figure 4B**).

**Figure 4.**
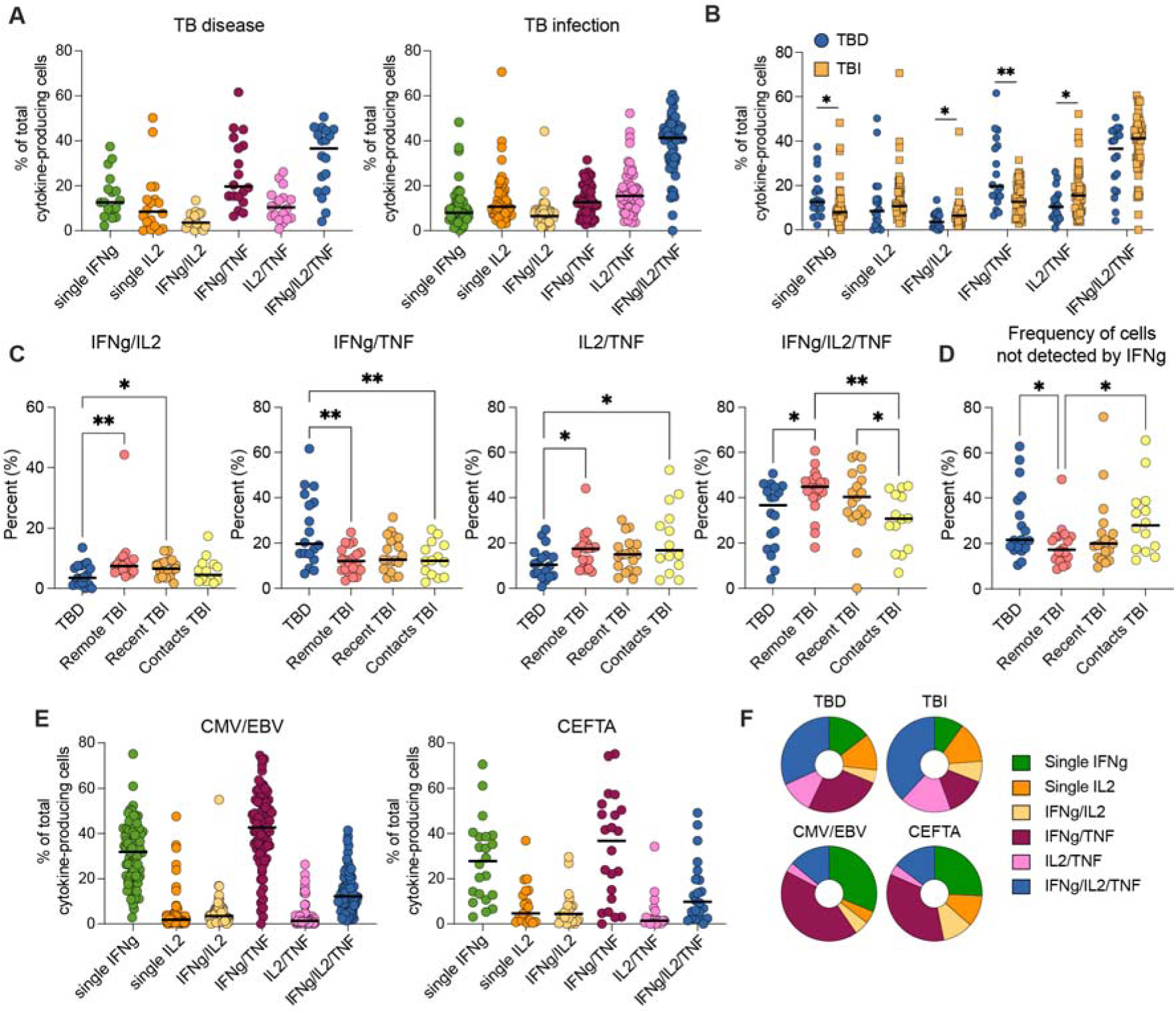
Cytokine combinations in response to pathogen specific peptide stimulation. (**A**) Frequency of all different cytokine combinations detected after stimulation with ESAT-6, CFP-10, EspC peptide pools in TBD (left panel) and TBI (right panel). (**B**) The frequency of single, double and triple cytokine-producing cells after stimulation with ESAT-6, CFP-10, EspC. Comparison between TBD (n=18) and TBI (n=53) Statistics was evaluated with the Mann-Whitney test. (**C-D**) Frequency of cytokine combinations between TBD (n=18), remote TBI (n=20), recent TBI (n=19), contacts (n=14). Statistics was evaluated using a Kruskal-Wallis test followed by uncorrected Dunn’s test. (**E**) Frequency of all different cytokine combinations detected after stimulation with peptide pools targeting T cells specific to CMV and EBV (left panel, n=80) or T cells specific to cytomegalovirus, Epstein-Barr virus, influenza, tetanus, and adenovirus (CEFTA) (right panel, n=22). (**F**) Donought plots indicating the proportion of different cytokine combinations for TBD, TBI, CMV/EBV, and CEFTA stimulated cells. Donors with < 25 total cytokine-producing cells were excluded. *p<0.05, **p<0.01. **Abbreviations:** TBD: TB disease; TBI: TB infection. CMV: Cytomegalovirus; EBV: Epstein-Barr virus.

We next investigated if there were differences associated with time since *Mtb* exposure within the TBI group. The group was divided into contacts (<6 months, n=14), recent (6-24 months, n=19), and remote (>24 months, n=20). Within the TBI group, contacts had significantly lower proportions of triple IFNγ/IL2/TNF+ cells compared with both recent and remote TBI, where proportions were gradually increasing (**Figure 4C**). The proportion of IFNγ/IL2+ cells was also higher in both recent and remote TBI when compared to TBD, a difference that was not seen when comparing contacts to TBD. Since the IGRA assay is based on IFNγ detection, we also calculated the proportion of cells not secreting IFNγ for each group, with both TBD and contacts having a larger proportion (median of 22% for TBD and 28% for contacts) compared with remote TBI (17%) (**Figure 4D**).

To understand if the *Mtb*-derived cytokine profile was specific to *Mtb* infection, we investigated the patterns of cytokine-secretion when cells were stimulated with peptides from other pathogens. PBMCs from the same donors were stimulated with peptide pools for CMV and EBV or for a subset of donors with peptide pools for CMV, EBV, Flu, Tetanus, and Adenovirus (CEFTA) (**Figure 4E**). These non-*Mtb* peptide pools induced similar cytokine secreting patterns, with the most frequent being double IFNγ/TNF+ cells followed by single IFNγ+ cells (**Figure 4E**). This contrasted with the overall *Mtb*-specific responses, which were dominated by IFNγ/IL2/TNF+ cells with few single IFNγ+ cells (**Figure 4F**).

### Adding IL2/TNF cytokine producing cells to total IFNγ increases detected SFU levels but does not improve assay accuracy

Since upon stimulation with *Mtb* antigens, 17-28% of SFUs did not contain IFNγ, we hypothesized that the addition of double IL2/TNF-producing cells could potentially increase the sensitivity for detecting *Mtb*-infection in TB infection and disease. We therefore combined the total IFNγ SFU with double IL2/TNF+ SFU (**Figure 5A**). This led to significantly increased SFU in all three groups (TBD, TBI, and controls). We then performed ROC analysis to evaluate if this would translate to better detection of TBD and TBI versus controls, but AUC remained unchanged in the combined total IFNγ and IL2/TNF in both comparisons (**Figure 5B**). Lastly, we assessed if adding IL2/TNF to total IFNγ could improve the percentage of detection positivity, but despite that SFU increased as an effect of the addition of IL2/TNF, it did not enable the detection of any additional donors compared with total IFNγ alone (**Figure 5C**). This indicates that all donors in this study that secreted detectable levels of IL2/TNF also produced detectable levels of IFNγ SFUs.

**Figure 5.**
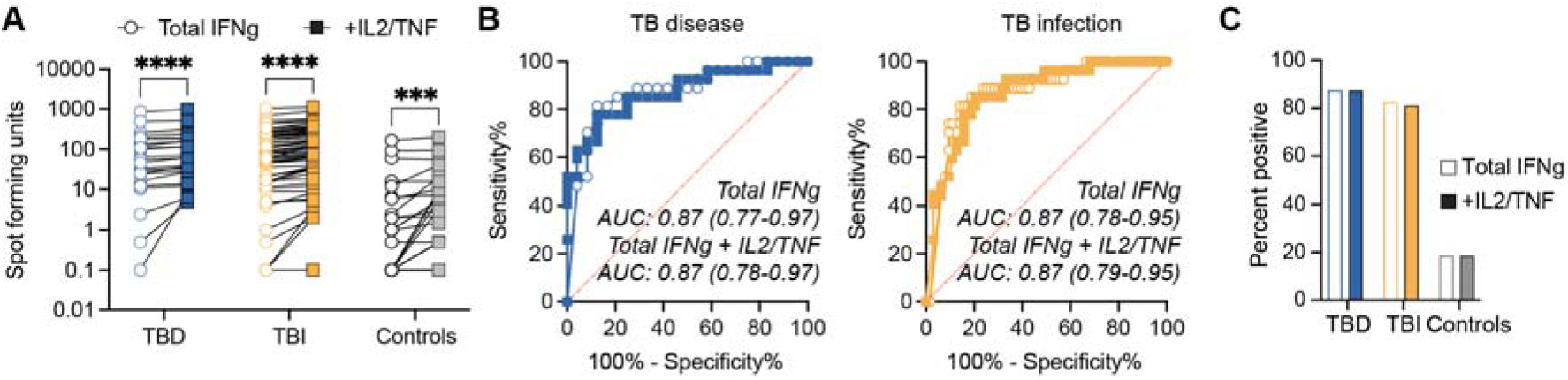
The addition of IL2/TNF-secreting cells to total IFNγ increase SFUs but not detection of TB infection or disease. (**A**) Number of (background-subtracted) SFU for total IFN*γ* (open circles) and total IFN*γ* with SFUs for TNF/IL2 added (filled boxes) for TBD (n=24), TBI (n=63), and IGRA negative controls (n=27). Statistics was evaluated using Wilcoxon matched pairs test with ***p<0.001 and ****p<0.0001. (**B**) AUROC curves for TB disease (left panel) and TB infection (right panel) versus IGRA negative controls comparing Total IFN*γ*-producing cells (open circles) with IFN*γ* + IL2/TNF (filled boxes). (**C**) Bar graphs indicating detection positivity for individuals with TBD (blue, n=24), TBI (orange, n=63) and IGRA negative controls (grey, n=27) based on [SFU(stimulated) – 2 x SFU(unstimulated) *≥* 5] for Total IFN*γ* (open bars) and IL2/TNF (filled bars). **Abbreviations:** SFU: spot forming units; TBD: TB disease; TBI: TB infection; IGRA: interferon gamma release assay. AUROC: area under the receiver operating characteristic.

### Quantitative differences in IFNγ, IL2, and TNF secretion associated with Mtb-infection status and time since Mtb exposure

In addition to counting the number of cytokine-secreting cells, we assessed the average relative spot volume as a measure of the amount of cytokine secreted. We calculated the average relative spot volume for each cytokine combination in samples with at least three *Mtb* peptide stimulated spots per well. Overall, in pooled TBD and TBI data, cells secreting only one single cytokine had the lowest spot volume, while cells producing all three cytokines had the largest spot volume, although not to a significant degree for IL2 (**Figure 6A**). Among intermediate producers, cells producing IFNγ/IL2 secreted more IFNγ compared with cells producing IFNγ/TNF but similar amounts of IL2 as those producing IL2/TNF, which in turn produced similar TNF levels as those secreted by cells producing IFNγ/TNF (**Figure 6A**).

**Figure 6.**
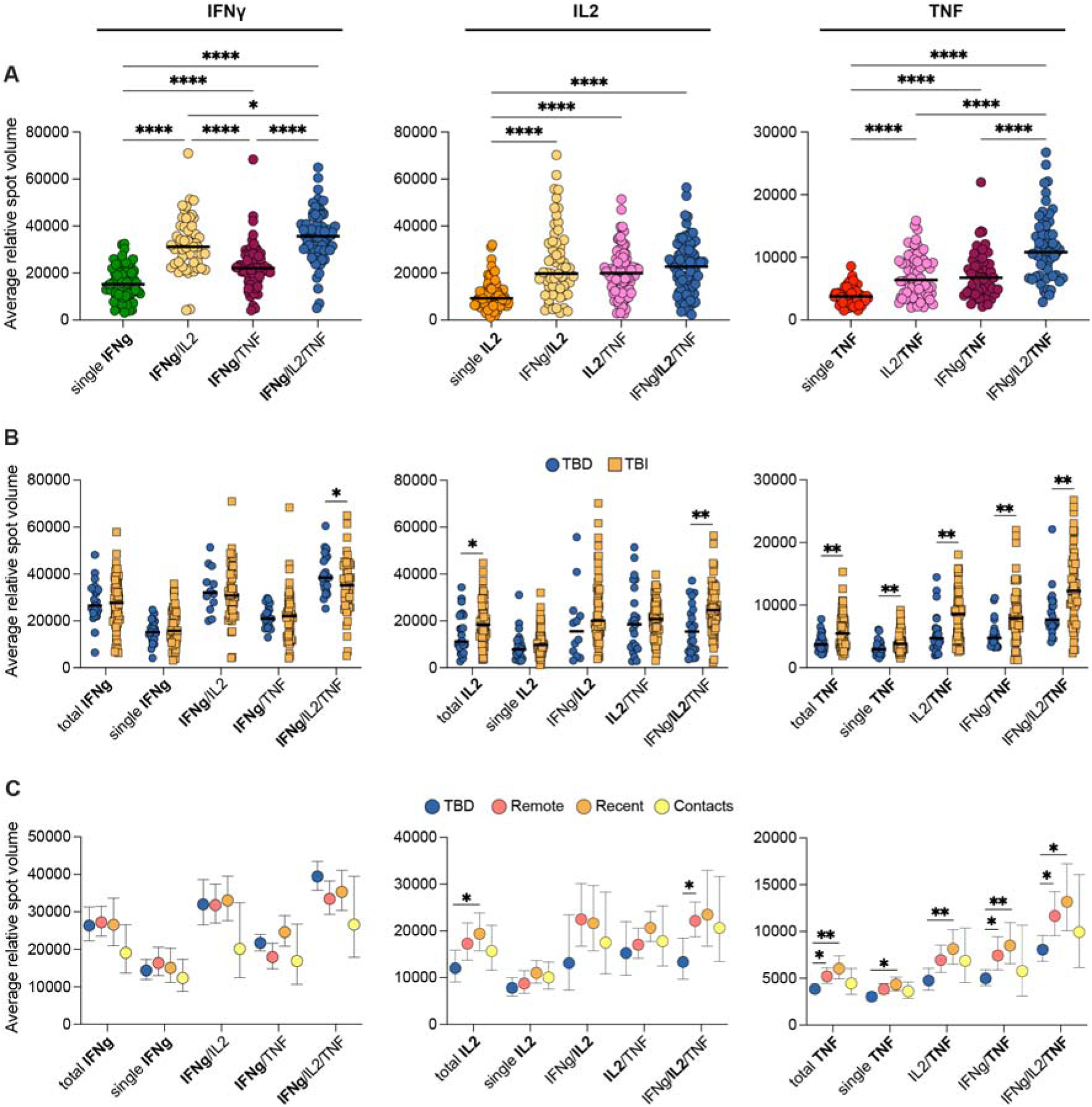
The magnitude of cytokine production is associated with cytokine combinations, Mtb infection status and time since Mtb exposure. Average relative spot volume for IFN*γ* (left), IL2 (middle), and TNF (right) spots in different cytokine combinations for (A) TBD and TBI donors combined (n=52-82). Statistics was evaluated using mixed-effects analysis followed by Tukey’s posttest on log10-transformed data. (**B**) TBD (n=12-24) and TBI (n=49-63) donors separated with statistics evaluated by Mann-Whitney tests. (**C**) TBD (blue, n=12-24) and TBI donors separated into remote (red, n=21-27), recent (orange, n=16-20), and contacts (yellow, n=12-16) with statistics evaluated using mixed-effects analysis followed by Tukey’s posttest on log10-transformed data. The cut-off to quantify spot volume in an individual sample was to have a spot number *≥* 3. Lines in A and B indicate median. Dots represent individual donors. Symbols and error bars in C represent geometric mean ± 95% CI. Only comparisons with p<0.05 are shown. *p<0.05, **p<0.01, ***p<0.001, ****p<0.0001. **Abbreviations:** TBD: TB disease; TBI: TB infection.

We next separated TBD and TBI and compared the relative spot volume between groups for each cytokine combination (**Figure 6B**). Triple IFNγ/IL2/TNF producing cells secreted on average more IFNγ in individuals with TBD compared with those with TBI. However, the converse was observed for IL2, where donors with TBI secreted overall more IL2 and especially among triple IFNγ/IL2/TNF producing cells. TNF on the other hand was secreted to a larger extent in all cytokine combinations in individuals with TBI (**Figure 6B**).

To further investigate if there were differences in the level of cytokine secretion associated with time since *Mtb* exposure, the TBI group was divided into contacts (<6 months, n=16), recent (6-24 months, n=20), and remote (>24 months, n=27) and spot volumes were compared between TBD and TBI subgroups for all cytokine combinations (**Figure 6C**). Although a clear trend was observed for reduced IFNγ secretion for TBI contacts, no significant differences could be determined. Consistent with the overall group data, individuals with TBD produced less IL2 compared with TBI. This was observed in all cytokine combinations, with statistical support for total IL2 and triple IFNγ/IL2/TNF secreting cells. In contrast, high TNF secretion was primarily associated with recent and remote TBI, while contacts secreted levels similar to individuals with TBD (**Figure 6C**).

## Discussion

In this study we evaluated a FluoroSpot assay with simultaneous detection of the three cytokines IFNγ, TNF and IL2 following overnight stimulation with peptides from the *Mtb* antigens ESAT-6, CFP-10 and EspC in individuals with TBD, TBI and IGRA negative controls. An inherent challenge when evaluating tests for TBI is that there is no reference method, as current tests are incapable of differentiating between persisting infection and immunological memory: therefore, sensitivity in TBD can be used as a surrogate measure and specificity is based on results in unexposed individuals from TB low-endemic settings [15].

Our results indicate that using triple IFNγ/IL2/TNF+ cells in TBD provided equal sensitivity to total IFNγ detection but had a higher specificity. Furthermore, the one IGRA negative individual with triple IFNγ/IL2/TNF+ cells above the set cut-off, was a person originating from a TB high-endemic country and a true TB infection could have gone undetected by a conventional IGRA-test. Considering that we stimulated cells with peptides from ESAT-6, CFP-10, and EspC, it is possible the primary reactivity in this donor was towards EspC, which is not included in the clinically used IGRA assay, indicating the value of extending the peptide pool used for assessing *Mtb-*specific T cells.

Triple IFNγ/IL2/TNF+ cells were the most frequently detected in both TB groups (TBD 37% and TBI 41% of SFUs), contrasting with responses to peptide pools specific to several viral infections, including EBV, CMV, adenovirus, and Influenza and the tetanus vaccine. Previous studies have reported partly conflicting findings with three studies based on flow cytometry suggesting that *Mtb*-specific triple IFNγ/IL2/TNF+ CD4+ T cells are more frequent in TBD than in TBI and contacts [48–50]. Methodological differences and diverse study populations however make direct comparisons difficult. Caccamo et al [49] measured T cell responses following stimulation with individual *Mtb* antigens (ESAT-6, Ag85B and 16kD) while we used a combined pool of *Mtb* antigens (ESAT-6/CFP-10/EspC). Additionally, our TBD cohort included both pulmonary and extrapulmonary TB while the referred studies all focused on pulmonary TB, with one including only individuals with smear positive pulmonary TB and the comparator group consisting of both IGRA positive and IGRA negative contacts [48], while another study also included non-culture confirmed pulmonary TB [50]. In this study, the IFNγ/IL2/TNF+ cells in TBI also produced less IFNγ compared with individuals with TBD, which could influence identification by flow cytometry depending on how the gate is set.

Somewhat obscuring the interpretation, a negative correlation between triple IFNγ/IL2/TNF+ cells and bacterial load in pulmonary TB has also been suggested, with levels inversely correlating to sputum smear grade and gradually increasing during TB treatment [51] while another study [49] concluded that following successful treatment of TBD, triple+ frequencies were the same as in controls. One possible explanation could be that when infection progresses from a controlled state (TBI) to a non-controlled state (TBD), the number of triple+ cells peak, before reducing in numbers and function due to immune exhaustion [52–55] resulting in loss of control of the disease and subsequent high bacterial load. As immunological control is regained during treatment, *Mtb* antigen levels decrease, and immune function may be restored, including a contraction of the triple+ T cell numbers. If that hypothesis is valid, it would mean that reduced numbers of triple+ cells could indicate complete clearance of *Mtb*. If so, this would be useful in the differentiation of TBI with or without persistent *Mtb* bacilli. Further prospective studies investigating polyfunctional *Mtb*-specific T cell responses after treatment completion would be important to address this question.

In addition to changes in the levels of cytokine-producing cells, there were also changes in the distribution of cytokine combinations. This could be associated with the *Mtb*-specific T cells being at varying stages of differentiation [56]. Consistent with this, we found that individuals with TBD had an increased proportion of cells producing more IFNγ and less IL2 compared with individuals with TBI. These results support previous findings suggesting a higher number of single IL2+ T cells in TBI, while triple IFNγ/IL2/TNF+ and single TNF+ T cells were more common in individuals with TBD than in exposed contacts [48]. This is also consistent with observations [49,57,58] where IL2/TNF+ and single IL2+ cells were associated with TBI. Additionally, Casey and al [58] examined IFNγ and IL2, using a dual FluoroSpot, and found a higher frequency of single IFNγ in TBD and a higher frequency of single IL2 and double IFNγ/IL2 secreting T-cells in TBI, coherent with our data.

IL2 is secreted from activated helper type 1 CD4 T cells (Th1), and serves several roles in T cell homeostasis, regulation of the function of effector T cells, and stimulating regulatory T cells differentiation [53,59]. We propose that in TBI and in particular recent TBI, IL2-producing central memory T (T_CM_) cells could be more frequent and play a role in mounting an early efficient immune response to *Mtb* infection. Conversely, upon losing control of the infection, T cells would differentiate toward effector memory (T_EM_) cells with reduced IL2 production and increased IFNγ and TNF secretion. In support of this hypothesis our data showed a higher proportion of IFNγ/TNF cytokine-producing cells in TBD compared to TBI. This data is further supported by findings by Arrigucci and co-workers [60], where they observed more IFNγ and TNF mRNA transcripts in individuals with TBD following *Mtb*-specific stimulation and higher ratios of T_EM_ to T_CM_. For our data, the balance between T_EM_ and T_CM_ cells could potentially explain the higher volume of secreted IFNγ in the triple+ cells in TBD, compared to more IL2 in the triple+ cells in TBI. Further indicating a change in IFNγ-producing cells associated with progression toward and development of TBD, previous studies of IFN-γ levels in QTF-plus and QTF-GIT have demonstrated a correlation between higher levels and the risk of progression to TBD [61,62], potentially indicating a loss of control and expanded differentiation to effector cells. Consistent with these patterns, another study showed that a dual IFNγ/TNF assay improved specificity (94%) for TBD vs TBI and non-TB compared to a single IFNγ assay without affecting sensitivity [57], suggesting that these changes in cytokine secretion can be explored for diagnostic purposes.

In summary, the FluoroSpot assay showed the highest AUC for TBD when using triple IFNγ/IL2/TNF+ cells, superior to only measuring IFNγ as in the current standard methods. By including multiple *Mtb*-specific responses (IL2/TNF together with total IFNγ or IFNγ/IL2/TNF) we could further increase SFU levels, indicating that more *Mtb* specific cells were detected. This could possibly have a clinical implication by improving detection of TBI in immunosuppressed individuals who may have reduced levels of circulating T cells, and where current methods might not reach the threshold for a positive test. In this specific patient group, at higher risk of progression to TBD, the corresponding lower specificity could be acceptable. However, additional studies are needed to explore performance in relevant patient cohorts. The positive predictive value of current IGRA-tests and TST for development of TBD are very low (at best around 5% [24]). This means that a more targeted approach for screening and treatment of TBI in larger populations is needed. Therefore, a potential benefit of a multiplex cytokine assay is that it would enable detection of a non-IFNγ or more IL2-dominated immune response in recent *Mtb* infection, of particular importance during contact tracing. Separate patterns related to recent and remote TBI would help identify which individuals are at higher risk of progression to TB disease and who therefore would have an increased benefit from TBI preventive treatment. Although FluoroSpot, similar to standard IGRAs, depends on laboratory facilities, we see a possible advantage over the single-IFNγ T-SPOT.TB assay. By learning more about the different cytokine expression patterns, this could aid in the development of other, less technically demanding, approaches.

### Limitations

In the assay described here, we cannot identify which cell types or differentiation stages of the cells that are producing the cytokines. For diagnostic purposes, this is of marginal relevance, however this limitation does not allow us to fully understand the biological processes involved. In our cohort we see a discrepancy between the clinical IGRA results and IFNγ-detection in the TBI group, where our assay displays a lower sensitivity. We believe this is due to a less streamlined process for handling of samples, although strict laboratory protocols were followed. In contrast to the clinical situation where tests are run the same day, our patient samples were stored in the freezer before the experiments were performed. Furthermore, the detection of positive study participants was also consistent with other studies done using thawed cells [63]. Additionally, even when comparing the different commercial IGRAs head-to-head, results are not fully congruent.

## Methods

### Ethical considerations

The study was registered and granted ethical permission from the Swedish Ethical Review Board, EPN-numbers 2013/1347-31/2 and 2019-05438. All study participants received written and verbal information and signed a written informed consent form prior to study inclusion. In relevant cases professional interpreter services were used.

### Study design

Prospective observational cohort study.

### Study participants

Adults (>18 years old) with TB disease or TB infection were prospectively recruited at convenience at the Infectious Disease Department, Karolinska University Hospital, Stockholm between May 2018 and November 2023. Eligible for the study were: 1) Patients with presumed or confirmed TB disease sampled within 1 week of anti-TB treatment initiation. TB disease was microbiologically verified via *Mtb* culture or *Mtb* PCR. 2) Individuals with TB infection defined as a positive IGRA result (QuantiFERON-TB Gold In-Tube [QFT-GIT] or QuantiFERON-TB Gold Plus [QFT-Plus]) mainly falling into three categories: i) close contacts of a TB index case; ii) screening of migrants from a TB high-endemic country (REF FoHM); iii) screening of patients in anticipation of immunosuppression or during pregnancy. TB disease was excluded by chest Xray, medical examination and if warranted mycobacterial sampling. 3) IGRA-negative controls consisted of medical students and staff without known or epidemiological risk of TB exposure (n=8), individuals tested via contact tracing (n=6), and patients investigated for TB disease (n=3) or infection (n=10) who upon follow-up had negative IGRA results and TB disease excluded.

### Data collection

Demographic, epidemiological, and clinical data for patients with TB disease and TB infection were extracted from patient charts. For all subjects this included information regarding previous history of or contact with individuals with TB disease, co-morbidities, current medication, and radiological, biochemical, and mycobacterial test results. In TB disease, the presence of typical TB-related symptoms (fever, night sweats, loss of appetite and weight loss) were noted, and patients were classified as pulmonary or extrapulmonary TB. Microbiological samples for mycobacterial analysis were collected independent of the study in accordance with clinical practice. All patient samples were analyzed at Karolinska University Hospital Laboratory with smear fluorescence microscopy (BX41, Olympus) for detection of acid-fast bacilli, PCR for detection of *Mtb* complex (BD MAX™ Systems or GeneXpert *MTB* Ultra), as well as mycobacterial culture in liquid (BACTEC™ MGIT™ 960) and solid (Löwenstein-Jensen) medium (inhouse). Positive microscopy results were graded as high/medium/low per standard laboratory protocol [64].

### Sample collection and processing

Venous blood samples were collected in EDTA vacutainer tubes (BD) and transferred to the laboratory for isolation of peripheral blood mononuclear cells (PBMCs) by density gradient centrifugation, using either SepMate tubes (Stemcell technologies) or Leucosep tubes (Thermo Fisher Scientific). The blood was transferred into 50 ml falcon tubes and diluted 1:1 with phosphate buffered saline (PBS) supplemented with 2% heat-inactivated fetal bovine serum (FBS) (Thermo Fisher Scientific). For the isolation with SepMate tubes, 15 ml Ficoll– Paque Plus (Cytiva) was added to 50 ml SepMate tubes and centrifuged for 1 minute at 1000 *g* in room temperature (RT). The diluted blood was poured into the SepMate tubes, above the insert, and centrifuged for 10 minutes at 1200 *g* RT, with the break on. The cells were then transferred to new 50 ml falcon tubes. For the Lecousep tubes, 15ml Ficoll-Plaque Plus was poured into 50 ml Leucosep tubes and centrifuged for 1 minute at 1000 *g*, RT. The diluted blood was poured into the Leucosep tube and subsequently centrifuged for 15 minutes at 800 *g,* RT, with the break turned off. The cell layer was collected using a Pasteur pipet and moved to a new 50 ml falcon tube. For both methods the cells were washed twice with 50 ml of PBS + 2% FBS (10 minutes centrifugation at 300 g, RT, brake at 5 out of 9) after collection. After the second wash the cells were stained with trypan blue and counted using a Countess II automatic cell counter (Invitrogen). The cells were centrifuged and resuspended at a concentration of 5 million cells/ml in freezing medium (FBS + 10% dimethyl sulfoxide) and moved to cryotubes. The tubes were placed inside a CoolCell and stored at – 80°C overnight. The following day the cells were transferred to liquid nitrogen for long-term storage.

### ELISpot and multiplex FluoroSpot

Peripheral blood mononuclear cells (PBMCs) were thawed in a 37 °C water bath, transferred to 15 ml falcon tubes and washed twice (300 *g* 5 min) in 15 ml complete media consisting of of RPMI-1640 supplemented with 10% fetal calf serum (FCS), 0.1 mM HEPES, 1% penicillin-streptomycin, and 1% L-glutamine (all from ThermoFisher). The cells were left to rest overnight in a 37 °C humidified incubator supplemented with 5 % CO_2_. On the second day the cells were filtered through a 70 μm nylon strainer, counted and resuspended to 5 × 10^6^ cells per mL in complete media. Pre-coated 96-well FluoroSpot Plus (IFNγ/IL2/TNF; Mabtech) or ELISpot Plus (IFNγ; Mabtech) plates were washed 5 times with 200 μl phosphate buffered saline (PBS, Merck) and blocked with 200 μl complete media for 30 min to 1 h at 37 °C. PBMCs were then seeded in then 96-well plates at 50 μl per well, corresponding to 2.5 × 10⁵ cells. An additional 50 μl of stimulation was added to each well giving a total volume of 100 μl per well. After addition of the stimulation, the plates were incubated overnight at 37 °C, 5% CO_2_.

The stimulations consisted of: 1) a mixed *Mtb* peptide pool containing peptides from EspC (#3623-1, n=23 peptides), CFP-10 (#3625-1, n=23 peptides), and ESAT-6 (#3624-1, n=21 peptides), 2) a mixed EBV (#3641-1, n=100 peptides) and CMV peptide pool (#3619-1, n=42 peptides), targeting both CD4+ and CD8+ T cells, 3) a CEFTA peptide pool (#3617-1, n=35 peptides) targeting CD4+ T cells. All peptide stimulations received co-stimulation with 0.1 μg/mL anti-CD28 (Mabtech). Unstimulated cells were used as a negative control. The final concentration of each peptide in the culture was 2 µg/mL.

After 16-24 hours in a humidified incubator with 5% CO₂, the plates were washed five times with 200 μl PBS, and 100 μl detection antibodies (either anti-IFNγ-biotin for ELISpot or a mix of anti-IFNγ-BAM, anti-IL2-biotin, and anti-TNF-WASP for FluoroSpot) were diluted in PBS + 0.1% BSA and added and incubated 2h at RT. After washing the plates five times, 100 μl fluorophore-conjugates (SA-ALP for ELISpot or anti-BAM-490, SA-550, and anti-WASP-640 for FluoroSpot) were diluted in PBS + 0.1% BSA and added for 1 h at RT. After additional washing five times, 50μl BCIP/NBT-plus was added followed by 5-10 min incubation before washing in tap water (for ELISpot) or 50 μl fluorescence enhancer was added for 15 min and then flicked out (for FluoroSpot) before plates were dried overnight at RT in the dark. Spot analysis was done on a Mabtech IRIS™ 2 FluoroSpot/ELISpot reader.

### Statistical evaluation

Statistics was evaluated using GraphPad prism version 10.4.2. Overall, due to the nature of the data distribution, spot forming units (SFU) were log10 transformed prior to calculating statistics using parametric tests or alternatively non-parametric tests were used. Data based on proportions or average relative spot volumes were calculated on non-transformed data. Two-way ANOVA or mixed-effects models were used followed by multiple tests with Tukey’s posttest for parametric data or Kruskal-Walls followed by multiple tests with (Mann-Whitney for unpaired and Dunn’s or Wilcoxon for paired) non-parametric data. P-values were indicated as *p<0.05, **p<0.01, ***p<0.001, ****p<0.0001.

## Acknowledgements

We thank the study participants for their contribution to the study. We also thank the study nurses Monica Modin, Jan Bellbrant, Anna Dahlberg, Lena Jansson, and Anna Löwhagen Welander for patient inclusion and Mabtech for providing FluoroSpot kits. This study was supported by grants from the Swedish Research Council (2021-03706 and 2023-01943), the Swedish Medical Association (SLS-934363 and SLS-960484), the Swedish Society for Medical Research (CG-24-0012-B), and the Heart-Lung Foundation (20220566 and 20230244) to CS. Grants from Swedish Research Council (2019-04663 and 2020-03602) and the Heart and Lung Foundation (20180386 and 20200194) to GK. Grants from The Swedish Physicians Against AIDS Research Fund (Fob2022-0012) to EF.

## Author contributions

CS, JB, BM, EF and GK designed the study. EF, VK, LG, MJ, JH, and JB included patients and collected clinical information. BM provided critical material and HG provided critical access to instruments. FF, VK, LG, MJ, JH, CSS, MG, and LK performed experiments. EF, VK, LG, MJ, and CS performed data analysis. CS and EF generated figures and tables. EF, MCN, GK, BM, JB, and CS interpreted the results. EF, CS, and GK provided funding. EF, JB, and CS provided project supervision and mentoring. EF, GK and CS wrote the initial draft with input from LK, FF, JB and MCN, and all authors contributed to revision and editing.

## Data availability statement

Data is available upon reasonable request to the corresponding author and pending confirmation of relevant permits.

## Additional information

Bartek Makower is an employee with the biotech company Mabtech AB. All other authors declare no competing financial interests.

